# Maternal and Infant Research Electronic Data Analysis (MIREDA): A protocol for creating a common data model for federated analysis of UK birth cohorts and the life course

**DOI:** 10.1101/2024.04.08.24305489

**Authors:** MJ Seaborne, HE Jones, N Cockburn, S Durbaba, TC Giles, A González-Izquierdo, A Hough, D Mason, A Mendez-Villalon, C. Sanchez-Soriano, C. Orton, D Ford, P Quinlan, K Nirantharakumar, L. Poston, RM Reynolds, G Santorelli, S Brophy

## Abstract

**Introduction:** Birth cohorts are valuable resources for studying early life, the determinants of health, disease, and development. They are essential for studying life course. Electronic cohorts are live, dynamic longitudinal cohorts using anonymised, routinely collected data. There is no selection bias through direct recruitment, but they are limited to health and administrative system data and may lack contextual information.

The MIREDA (Maternal and Infant Research Electronic Data Analysis) partnership creates a UK-wide birth cohort by aligning existing electronic birth cohorts to have the same structure, content, and vocabularies, enabling UK-wide federated analyses.

**Objectives:**

1. Create a core dynamic, live UK-wide electronic birth cohort with approximately 100,000 new births per year using a common data model (CDM).
2. Provide data linkage and automation for long-term follow up of births from MuM-PreDiCT and the ‘Born in’ initiatives of Bradford, Wales, Scotland, and South London for comparable analyses.

**Methods:** We will establish core data content and collate linkable data. Use a suite of extraction, transformation, and load (ETL) tools will be used to transform the data for each birth cohort into the CDM. Transformed datasets will remain within each cohort’s trusted research environment (TRE). Metadata will be uploaded for the public to the <u>Health Data Research (HDRUK) Innovation Gateway</u>. We will develop a single online data access request for researchers. A cohort profile will be developed for researchers to reference the resource.

**Ethics:** Each cohort has approval from their TRE through compliance with their project application processes and information governance.

**Dissemination:** We will engage with researchers in the field to promote our resource through partnership networking, publication, research collaborations, conferences, social media, and marketing communications strategies.

## Introduction

Rapid socio-economic changes including the recent increased cost of living in the UK are contributing to widening inequalities which will impact population health (1). This is coupled with the increased burden to the NHS, social care services and education disruption after the pandemic (2). These changes are likely to adversely affect maternal, infant and child health outcomes as there are strong negative associations with deprivation and poor diet (3), unhealthy maternal BMI (4,5), shorter intervals between pregnancies (6), pregnancy in adolescence (7), negative health behaviours (8), and conditions such as anaemia (9). These alone can be responsible for causal sequences which may result in deteriorating maternal, infant and child health and wellbeing, and can be damaging to development and future health and prosperity of offspring through their life course (10). There is a clear need for continued monitoring and evaluation of interventions for preventative strategies that reduce the risks of poor health and wellbeing in these population groups throughout their life course.

Although new technologies and treatments continue to improve population health, there is a greater need for informed preventative action that reduces the burden on healthcare resources (11). Measures to optimise maternal, infant and child health during the perinatal period and beyond will inevitably confer substantial benefits in the prevention of ill-health.

Traditionally, data from birth cohorts have been valuable resources for studying early life and the determinants of health, disease and development (12). They have played an essential role in profiling the life course of a child from conception through to all other stages of development.

Large birth cohorts have historically contributed to life course approaches to disease prevention and public health, they are often through direct recruitment, and are usually specific to a country, region, or other measure of locality. Directly recruited cohorts can limit representation of the target population because of selective inclusion. They are often limited by the period for which the cohort is defined and may not represent those born outside the cohort recruitment window (12–14).

Life course studies from birth are invariably complex as there are multiple and often interacting factors to consider which may influence outcomes. As such, the type and content of data collected is extensive and may include maternal characteristics (age, parity, gravida, ethnicity, deprivation scores, medical history, etc), pregnancy complications - both physical and mental health, birth outcomes (gestational age, birth weight, level of care required, etc), and other longitudinal data which relates to social care, developmental milestones, education, household composition, etc.

We are living in a new era with increasing access to vast amounts of health data and with growing analytical expertise, to interpret and disseminate meaningful insights to the health of our nations. This provides an exciting new capacity to further our research into the life course. Much work was done during the COVID-19 pandemic to bring together large amounts of health data to monitor its effects, but significant issues emerged when comparing routinely collected data across different nations and regions within the UK. These were compounded by a lack of standardisation of clinical coding systems, terminology, definitions, formats, difference in type and/or quantity of data collected, and governance relating to access to data (15). These differences were a barrier to accessibility of easily comparable data.

The evolution of data science has led to the development of accessible routinely collected anonymised data for large populations in trusted research environments (TREs) that can be linked to many different data sources. They provide rich, detailed information and remove obstacles such as the need to recruit, and so provide a means for which a cohort representative of that population can grow over time and the data remains relevant in the future.

In the UK, there is currently no central repository to access birth data for all four nations which is suitably anonymised and capable of linking to the plethora of data sources that help define the life course of each birth. National core datasets exist, but they are used for more general statistical output and without the granularity of data available through data linkage. There are several electronic birth cohorts which use different approaches to compiling routinely collected data for analysing population level data and the health life course. Each has similar data but with additional linkable datasets which lend them different strengths (16–20). They have arisen, in parallel, with improved relative ease of access, the removal of some research barriers and the need for analyses within specific populations which cannot necessarily be extrapolated from other, dissimilar populations. Harmonisation of these cohorts is a goal that aligns with the FAIR Guiding Principles for scientific data management and stewardship (21). The principles aim to establish comparability and consistency of analyses through a widely accepted standard. Achieving this would result in a network of cohorts gathering uniform core data, augmented with additional datasets that could be linked to extend the breadth and depth of research possibilities. This harmonisation effort would not only improve the findability and accessibility of birth data across the four nations but also enhance the interoperability and reusability of the data for diverse research needs, thus fulfilling the FAIR criteria and significantly advancing the field of life course health research.

Using the established standards of the Observational Medical Outcomes Partnership (OMOP) Common Data Model (CDM) could help achieve all this. This approach is used to standardise both the structure and content (terminologies, vocabularies, and coding schemes) of observational data and has been used successfully in other health data analyses to produce reliable evidence and enable comparisons. The OMOP CDM contains 37 standardised tables relating to: clinical data and vocabularies. It also contains additional domains relating to health systems, health economics, derived elements, and metadata (16).

The Mother and Infant Research Electronic Data Analysis (MIREDA) partnership will facilitate a platform in which UK wide, dynamic live birth cohorts will be collated into common core datasets using the OMOP CDM. The core data resource will initially contain the harmonised data for 1,000,000 live births and will grow by approximately 100,000 births per year, with long-term follow up using routine data. It will contain each respective cohort’s linkable data from public health, neonatal health, imaging, primary and secondary care and will be a central access point for longitudinal birth cohort federated analyses; federated analyses with the purpose of improving infant and maternal health across the UK by comparing differences in policies and practises.

The cohort will be facilitated by Health Data Research UK (HDR-UK) and access will be governed by their TRE principles and best practices (22). Accredited researchers wishing to undertake research with the harmonised OMOP cohorts will apply for access using a single, centrally approved HDR-UK application.

The metadata for the MIREDA resource will be prepared and made accessible from the <u>Health Data Research (HDRUK) Innovation Gateway</u> using a standardised request form.

The anonymised birth cohorts included are: Born in Wales (BiW) (17), Born in Scotland (BiS) (20), Born in Bradford (BiBBS and BiB4ALL) (23–25), the early Life data Cross-Linkage in Research (eLIXIR) partnership (Born in South London) (18), and Multimorbidity in Pregnancy: Determinant, Consequences, Clusters and Trajectories (MuM-PreDiCT) (19).

Additional linkable data, where available for each cohort will be provided and used to enrich datasets beyond their core content.

This phase of the project will run for one year from June 2023.

## Methods

### Study design

The MIREDA partnership will bring together standardised, harmonised birth cohorts relating to mother, baby, and child in a federated common data framework. This will include structured definitions, descriptions and documentation of the data using the OMOP common data model (CDM), a comprehensive data dictionary, a data quality plan, and data governance policy. The data will be available for creating a sustainable and reactive health system to inform public health decisions and policy.

The project unites the cohorts from BiW, eLIXIR (BiSL), BiBBS/BiB4ALL, BiS, and MuM-PreDiCT, encompassing data from all four UK nations. The initial combined cohort will represent approximately 350,000 live births from the first four cohorts. The share from each of these cohorts is apportioned as 82%, 13%, 4%, and 1% respectively. MuM-PreDiCT will represent approximately eight million births across the UK.

We estimate that the combined cohorts will amass an additional 100,000 births per year.

All live births on or after January 1^st^, 2014, will be identified from routine data for BiW and represent the earliest births in the collection. Others will contribute birth data from the start of their respective cohorts.

A suite of extraction, transformation and load (ETL) tools (WhiteRabbit (26), Convenient and Reusable Rapid OMOP Transformer (CaRROT) Mapper (27), and CaRROT-CDM (28) will be used to restructure concepts into the OMOP CDM of standard tables, fields and vocabularies.

### Data storage

Each electronic birth cohort with person-level data including outcomes and exposures will remain inside their own trusted research environment (TRE) and will be subject to all the established rules of their TREs.

### Core data: Cohort data alignment

The MIREDA partnership will collaborate to review the data accessible to each birth cohort and establish a consensus for their common data. This will be used to form a mutually agreed core dataset to be implemented for each birth cohort (Table *1*).

**Table 1.** Core data for birth cohort dataset modules.

| Dataset modules | Data | Additional information |
| --- | --- | --- |
| Maternal socio-demographics | Week/Date of birth |  |
|  | Ethnicity |  |
|  | Deprivation Quintile | By locality at start of pregnancy |
|  | Rural/urban | By locality at start of pregnancy |
|  | Education |  |
|  | Qualifications |  |
|  | Occupation |  |
| Baby socio-demographics | Week/Date of birth |  |
|  | Sex |  |
|  | Ethnicity |  |
|  | Deprivation quintile | By locality at birth and school age |
|  | Rural/urban residence | By locality at birth and school age |
|  | Disability codes |  |
| Pregnancy | Date of antenatal visit |  |
|  | Maternal age | At booking visit/conception |
|  | Maternal smoking status |  |
|  | Maternal Height |  |
|  | Maternal Weight |  |
|  | Maternal BMI |  |
|  | Existing diagnosis codes | Comorbidities existing at booking visit |
|  | New event type | New events since previous visit e.g. diagnosis, procedure, medicines, etc. |
|  | New event code |  |
|  | New event date |  |
|  | New event source | Event sources e.g. GP/hospital. |
|  | Data of expected Delivery |  |
|  | Folic acid supplements indicator |  |
|  | Domestic violence indicator |  |
|  | WHOOLEY score | Or other depression screening indicators. |
|  | Gravidity |  |
|  | Parity |  |
|  | Previous C-Section indicator |  |
|  | Date of previous C-section |  |
|  | Previous preterm birth indicator |  |
|  | Date of previous preterm birth |  |
|  | Gestational age | At visit. |
| Post-birth (maternal) | Postnatal depression indicator |  |
|  | Event type | Events within 10 weeks of birth date; may include: diagnosis, procedure, medication, etc. |
|  | Event code |  |
|  | Event date |  |
|  | Event source | Event sources e.g. GP/hospital. |
| Birth | Date of delivery |  |
|  | Gestational age | At birth. |
|  | Birth weight |  |
|  | Event type | Events from birth to 28 days; may include: diagnosis, procedure, medication, etc. |
|  | Event code |  |
|  | Event date |  |
|  | Event source | Event sources e.g. GP/hospital. |
|  | Mode of delivery | E.g. c-section, forceps, vaginal, etc. |
| Infancy | Event type | As above but for events from 29 days to school age |
|  | Event code |  |
|  | Event date |  |
|  | Event source |  |
|  | Neonatal admission indicator |  |
|  | Neonatal event indicator |  |
|  | Breastfeeding indicator | Breastfeeding, mixed or formula indicator |
|  | Breastfeeding duration |  |
| School age | Event type | As above but for events from school start age to 12 years. |
|  | Event code |  |
|  | Event date |  |
|  | Event source |  |

Details of linkable data for each cohort will also be provided in later work Table 2.

**Table 2.**
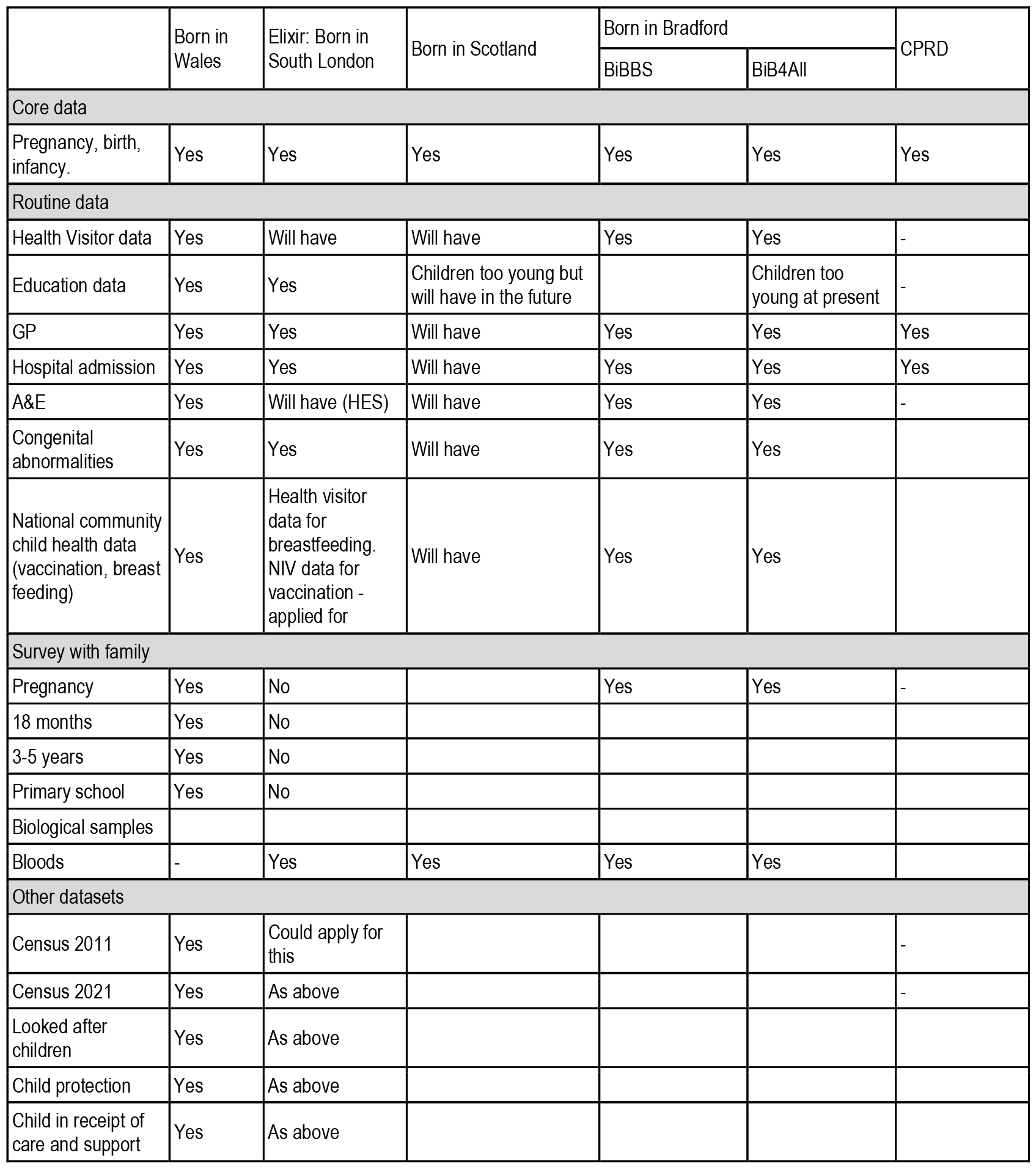
Linkable datasets.

|  | Born in Wales | Elixir: Born in South London | Born in Scotland | Born in Bradford |  | CPRD |
| --- | --- | --- | --- | --- | --- | --- |
|  |  |  |  | BiBBS | BiB4All |  |
| Core data |  |  |  |  |  |  |
| Pregnancy, birth, infancy. | Yes | Yes | Yes | Yes | Yes | Yes |
| Routine data |  |  |  |  |  |  |
| Health Visitor data | Yes | Will have | Will have | Yes | Yes | - |
| Education data | Yes | Yes | Children too young but will have in the future |  | Children too young at present | - |
| GP | Yes | Yes | Will have | Yes | Yes | Yes |
| Hospital admission | Yes | Yes | Will have | Yes | Yes | Yes |
| A&E | Yes | Will have (HES) | Will have | Yes | Yes | - |
| Congenital abnormalities | Yes | Yes | Will have | Yes | Yes |  |
| National community child health data (vaccination, breast feeding) | Yes | Health visitor data for breastfeeding. NIV data for vaccination - applied for | Will have | Yes | Yes |  |
| Survey with family |  |  |  |  |  |  |
| Pregnancy | Yes | No |  | Yes | Yes | - |
| 18 months | Yes | No |  |  |  |  |
| 3-5 years | Yes | No |  |  |  |  |
| Primary school | Yes | No |  |  |  |  |
| Biological samples |  |  |  |  |  |  |
| Bloods | - | Yes | Yes | Yes | Yes |  |
| Other datasets |  |  |  |  |  |  |
| Census 2011 | Yes | Could apply for this |  |  |  | - |
| Census 2021 | Yes | As above |  |  |  | - |
| Looked after children | Yes | As above |  |  |  |  |
| Child protection | Yes | As above |  |  |  |  |
| Child in receipt of care and support | Yes | As above |  |  |  |  |

The agreed core datasets will be created within each cohort. They will include all live births from 2014 or earliest year available after 2014.

Where available, core data will relate to both mother and baby with socio-demographic data such as date of birth, sex of baby, maternal education/qualifications, ethnicity, deprivation level, relationship status, and geographical location type (urban or rural). Other data will be divided into the following modules: pregnancy, maternal ten weeks post-birth, birth, infancy, and school age (Table 1).

### Data extraction and transformation

When data wrangling is complete and the core datasets are assembled, each site will use WhiteRabbit (24,25) software to scan them and produce scan reports. The reports will be anonymised and contain detailed information about tables, fields, and values that appear in fields. The anonymous summary reports will be extracted from the TREs for mapping and to create transformation rules.

The transformation rules will be imported into each TRE as a .json file and will be used by the locally installed CaRROT-CDM tool to transform the core datasets into OMOP CDM.

### Data access

Data access will be granted by central request via the <u>Health Data Research (HDRUK) Innovation Gateway</u>. Here the MIREDA collection will be available. It will detail the metadata for each cohort and their linkable datasets. We will adapt the current HDRUK data access request (DAR) form to satisfy each TRE’s requirements and create a single access request form for all MIREDA cohorts. No person-level data will leave any of the TREs. Instead, access will be used to create anonymised, aggregated tables and results of statistical analyses within each TRE. To do so, common R scripts will be developed and sent from a central hub to each TRE (step 1), where they will be implemented locally to produce the output required and will be inspected by local analysts (step 2) before transferring to the central hub (step 3) for synthesis and further analysis (step 4), Figure 1.

**Figure 1.**
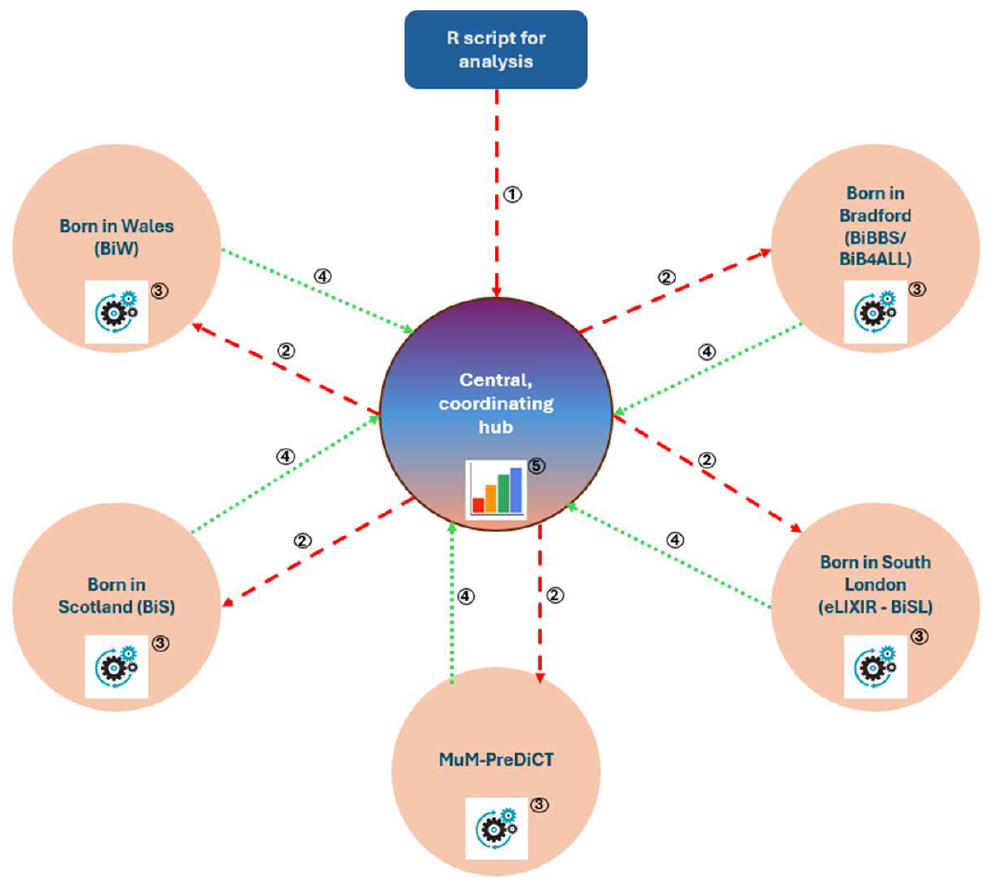
Federated analysis infrastructure. The numbers in the figure describe the following steps: 1. Analysis R script sent to central data hub (TRE) with specifications for the data model and analyses. 2. These are relayed to each birth cohort’s TRE. 3. R scripts are run on each TRE’s own system to create anonymised data tables and statistics. 4. These are sent back to the central hub for analysis. 5. Central hub compiles and analyses the data.

## Discussion

This protocol outlines the construction of a federated UK longitudinal birth cohort, leveraging routine data from UK healthcare providers, structured in a multi-modular format encompassing various stages from pregnancy to early childhood. The adoption of the OMOP CDM for this initiative is pivotal, ensuring that the FAIR principles are embedded within the data management framework. By defining core variables and incorporating supplementary data from diverse sources, the protocol facilitates the enhancement and contextualisation of the core datasets. Utilising the OMOP CDM allows for the standardisation of data, enabling uniform analyses across cohorts and fostering the use of a single, centralised coding strategy across different Trusted Research Environments (TREs). Consequently, this standardised approach permits the execution of a single analysis script across multiple cohorts, obviating the necessity for distinct scripts for each dataset.

The harmonised datasets created under this common data model will empower network studies, allowing for integrated research across different centres while ensuring that TREs maintain control over their original data. This is crucial for upholding the security protocols and governance standards of each TRE. Moreover, the central management of access requests via the HDR-UK gateway ensures a consistent application process that meets the specific requirements of each TRE site. This centralised system aligns with the FAIR principles by providing a transparent, efficient, and universally applicable approach to data access, thereby advancing the integrity and utility of research conducted on these valuable cohorts.

### Strengths and limitations of this study

It is a strength that this methodology uses an established standard for data harmonisation and standardisation through use of the OMOP common data model to create comparable core datasets for research.

The process will establish a comprehensive, UK-wide database which consolidates clinical data from maternity, neonatal, child health and education records. It will also be enriched by quantitative and qualitative results from surveys conducted by Born in Wales and Born in Bradford providing context and insight into detail not represented by routinely collected data.

The limitation of using routine and administrative data is that data may be missing or prone to errors. The data could lack context and there is a potential for loss of data pertaining to individuals who relocate outside of a cohort’s remit during pregnancy or after the child’s birth.

### Future directions

As the OMOP CDM has been used as an accepted standard internationally, future development will include working with other cohorts, and other countries for international research comparisons and insights. MIREDA intends to expand the OMOP standard to other non-healthcare data e.g. releated social and community data, and to apply a pseudo-OMOP style solution to inclusion. Differences between cohorts’ foci will be exploited for extrapolation to others where the same data or depth of data is not available. MIREDA aims to use pooled cohort data to address rare conditions. We seek to holistically prevent and reduce risk factors that adversely affect child health and wellbeing. We will work with HDR-UK to develop training and education materials, and to build a library of commonly used coding scripts for rapid output analysis.

## Conclusion

OMOP CDM is used internationally as an accepted standard for healthcare data and thus has a well-developed training and support infrastructure, so it is admirably suited to standardising data for collaborative use and to build stronger networks with improved insight into specific research topics. It allows for rapid, repeatable comparison of data between different regions, countries, and other localities. As there are many software tools and a network of experts available to support such work via the <u>Observational Health Data Sciences and Informatics (OHDSI)</u> group it is a relatively easy methodology to adopt and assimilate into existing TRE environments so that existing policies, procedures and standards can be maintained.

## Acknowledgements

This work was supported by an MRC Partnership Grant [MR/X02055X/1], MatCHNet pump-priming [U20005/302873] and an MRC Programme Grant [MR/X009742/1].

## Ethics Statement

Access to data is granted according to the information governance requirements of each TRE. The Data Protection Act 2018 is not applicable to anonymised data and the OMOP CDM will be anonymised and provide aggregated data and statistics only. Each TRE has ethical approval for its operation and use, thus no additional ethical approval was required beyond the standard project approval by official channels.

## Dissemination

The MIREDA partnership’s activities will actively engage, communicate, and disseminate this new data resource among researchers in maternal and infant health, the target research community. We will promote the resources, tools, collaborative expertise and infrastructure through publication, research collaborations, conferences, social media/marketing communications strategies.

MIREDA will present at meetings and events held by relevant health, and health data, research organisations. Workshops at targeted conferences will promote the linked data to encourage applications for grants and research collaborations.

Face-to-face workshops will present research with MIREDA data resources. Recordings will be uploaded as online webinars. Key groups invited will include policymakers, NHS healthcare professionals from maternity, and industry stakeholders.

MOOCs (massive open online courses) will be developed to improve skills and expertise in maternal and infant linked data analysis. Regular blogs and podcasts will discuss data use.

We will work with international partners to raise awareness of MIREDA for research collaborations.

MIREDA information will be hosted on the websites of MIREDA partners, and resources will be promoted through ADR Digital Insights.

Training will be provided in association with HDRUK training group, ADR training and NIHR to develop capacity and expertise. Pump priming funding will enable early career researchers to utilise the data.

## Patient and public involvement

Patients and/or the public were involved in the design, or conduct, or reporting, or dissemination plans of this research. We follow the Co-production of Research and Strategy (CORDS) standard operating procedure and the UK standards for PPI involvement and National Institute for Health Research (NIHR) guidance from INVOLVE (29). Records of PPI activity are maintained using the Public Involvement in Research Impact Toolkit (PIRIT) (30). Each cohort and co-connect works with its own PPI groups under different names/guises to contribute to each step of this process.

## Data availability statement

Data will be available upon reasonable request through the <u>Health Data Research (HDRUK) Innovation Gateway</u>.

## Abbreviations

MIREDA: Maternal and Infant Research Electronic Data Analysis
OMOP: Observational Medical Outcomes Partnership
CDM: Common Data Model
TRE: Trusted Research Environment
BiW: Born in Wales
eLIXIR: early Life data Cross-Linkage in Research
BiSL: Born in South London
BiB4All: Local name for BaBi Bradford
BaBi Bradford: Born and Bred in Bradford
BiBBS: Born in Bradford Birth Study
BiS: Born in Scotland
MuM-PreDiCT: Multimorbidity in Pregnancy: Determinant, Consequences, Clusters and Trajectories
ETL: Extraction, Transformation, and Load
HDR-UK: Health Data Research UK
GP: General Practitioner
CPRD: Clinical Practice Research Datalink
HES: Hospital Episode Statistics
NIV: National Immunisation Vaccine
DAR: Data Access Request
MOOC: Massive Open Online Courses
WHO: World Health Organization
NIHR: National Institute for Health Research
OHDSI: Observational Health Data Sciences and Informatics
PPI: Patient and Public Involvement
FAIR: Findable, Accessible, Interoperable, Reusable

## Full URLs contained within manuscript

Health Data Research Innovation Gateway https://www.healthdatagateway.org/

Observational Health Data Sciences and Informatics (OHDSI) https://www.ohdsi.org/

## References

1. Broadbent P, Thomson R, Kopasker D, Mccartney G, Meier P, Richiardi M, et al. The public health implications of the cost-of-living crisis: outlining mechanisms and modelling consequences. Lancet Reg Health - Eur. 2023;27:100585.

2. Fears R, Gillett W, Haines A, Norton M, Meulen V ter. Post-pandemic recovery: use of scientific advice to achieve social equity, planetary health, and economic benefits. Lancet Planet Health. 2020;4:e383–4.

3. Haggarty P, Campbell DM, Duthie S, Andrews K, Hoad G, Piyathilake C, et al. Diet and deprivation in pregnancy. Br J Nutr. 2009 Nov;102(10):1487–97.

4. Black RE, Allen LH, Bhutta ZA, Caulfield LE, Onis M de, Ezzati M, et al. Maternal and child undernutrition: global and regional exposures and health consequences. Lancet Lond Engl. 2008;371(9608):243–60.

5. Nurul-Farehah S, Rohana AJ. Maternal obesity and its determinants: A neglected issue? Malays Fam Physician Off J Acad Fam Physicians Malays. 2020;15(2):34.

6. Schummers L, Hutcheon JA, Hernandez-Diaz S, Williams PL, Hacker MR, Vanderweele TJ, et al. Association of Short Interpregnancy Interval With Pregnancy Outcomes According to Maternal Age. JAMA Intern Med. 2018 Dec;178(12):1661–70.

7. Chen XK, Wen SW, Fleming N, Demissie K, Rhoads GG, Walker M. Teenage pregnancy and adverse birth outcomes: a large population based retrospective cohort study. Int J Epidemiol. 2007 Apr;36(2):368–73.

8. Smoking and pregnancy patient information leaflet | RCOG [Internet]. Available from: https://www.rcog.org.uk/for-the-public/browse-our-patient-information/smoking-and-pregnancy-patient-information-leaflet/

9. Nair M, Churchill D, Robinson S, Nelson-Piercy C, Stanworth SJ, Knight M. Association between maternal haemoglobin and stillbirth: a cohort study among a multi-ethnic population in England. Br J Haematol. 2017 Dec;179(5):829–37.

10. Mikkelsen B, Williams J, Rakovac I, Wickramasinghe K, Hennis A, Shin HR, et al. Life course approach to prevention and control of non-communicable diseases. BMJ [Internet]. 2019 Jan;364. Available from: https://www.bmj.com/content/364/bmj.l257 https://www.bmj.com/content/364/bmj.l257.abstract

11. Hochlaf D, Quilter-Pinner H, Kibasi T. THE CASE FOR A NEW APPROACH TO PUBLIC HEALTH AND PREVENTION The progressive policy think tank. 2019; Available from: https://www.ippr.org

12. O’connor M, Spry E, Patton G, Moreno-Betancur M, Arnup S, Downes M, et al. Better together: Advancing life course research through multi-cohort analytic approaches. Adv Life Course Res [Internet]. 2022;53. Available from: http://creativecommons.org/licenses/by/4.0/

13. Downs JM, Ford T, Stewart R, Epstein S, Shetty H, Little R, et al. An approach to linking education, social care and electronic health records for children and young people in South London: a linkage study of child and adolescent mental health service data. Available from: http://bmjopen.bmj.com/

14. Overy C, Reynolds LA, Tansey EM, Group H of BR. History of the Avon longitudinal study of parents and children (ALSPAC), c. 1980-2000: the transcript of a Witness Seminar held by the History of Modern Biomedicine Research Group, Queen Mary, University of London, on 24 May 2011. 2012;122.

15. Data Standardization – OHDSI [Internet]. Available from: https://www.ohdsi.org/data-standardization/

16. Wright J, Small N, Raynor P, Tuffnell D, Bhopal R, Cameron N, et al. Cohort Profile: The Born in Bradford multi-ethnic family cohort study. Int J Epidemiol. 2013;42:978–91.

17. Jones H, Seaborne MJ, Kennedy NL, James M, Dredge S, Bandyopadhyay A, et al. Cohort profile: Born in Wales—a birth cohort with maternity, parental and child data linkage for life course research in Wales, UK. BMJ Open. 2024 Jan;14(1):e076711.

18. Carson LE, Azmi B, Jewell A, Taylor CL, Flynn A, Gill C, et al. Cohort profile: the eLIXIR Partnership-a maternity-child data linkage for life course research in South London, UK. BMJ Open. 2020;10:39583.

19. Lee SI, Eastwood KA, Moss N, Azcoaga-Lorenzo A, Subramanian A, Anand A, et al. Protocol for the development of a core outcome set for studies of pregnant women with pre-existing multimorbidity. BMJ Open. 2021 Oct;11(10):e044919.

20. Born in Scotland | The University of Edinburgh [Internet]. Available from: https://www.ed.ac.uk/cardiovascular-science/born-in-scotland

21. Wilkinson MD, Dumontier M, Aalbersberg IjJ, Appleton G, Axton M, Baak A, et al. The FAIR Guiding Principles for scientific data management and stewardship. Sci Data. 2016 Mar 15;3(1):160018.

22. Alliance UHDR, NHSX. Building Trusted Research Environments - Principles and Best Practices; Towards TRE ecosystems. 2021 Dec; Available from: https://zenodo.org/record/5767586

23. Dickerson J, Bridges S, Willan K, Kelly B, Moss RH, Lister J, et al. Born in Bradford’s Better Start (BiBBS) interventional birth cohort study: Interim cohort profile. Wellcome Open Res. 2022 Oct;7:244.

24. BiB4ALL [Internet]. Born In Bradford. [cited 2024 Feb 13]. Available from: https://borninbradford.nhs.uk/what-we-do/cohort-studies/born-in-bradford/

25. Cohort studies: BiB4ALL [Internet]. 2024. Available from: https://borninbradford.nhs.uk/what-we-do/cohort-studies/born-in-bradford/

26. WhiteRabbit for ETL design – OHDSI [Internet]. Available from: https://www.ohdsi.org/analytic-tools/whiterabbit-for-etl-design/

27. About - CaRROT-Mapper [Internet]. Available from: https://hdruk.github.io/CaRROT-Docs/CaRROT-Mapper/about/

28. About - CaRROT-CDM [Internet]. Available from: https://hdruk.github.io/CaRROT-Docs/CaRROT-CDM/About/

29. INVOLVE. Guidance on co-producing a research project [Internet]. INVOLVE; 2018. p. 1–20. Available from: https://www.invo.org.uk/posttypepublication/guidance-on-co-producing-a-research-project/

30. Public Involvement in Research Impact Toolkit (PIRIT) - Marie Curie Research Centre - Cardiff University [Internet]. Available from: https://www.cardiff.ac.uk/marie-curie-research-centre/patient-and-public-involvement/public-involvement-in-research-impact-toolkit-pirit

